# Empowering Personalized Pharmacogenomics with Generative AI Solutions

**DOI:** 10.1101/2024.02.21.24302946

**Authors:** Mullai Murugan, Bo Yuan, Eric Venner, Christie M. Ballantyne, Katherine M. Robinson, James C. Coons, Liwen Wang, Philip E. Empey, Richard A. Gibbs

## Abstract

**Objective:** This study evaluates an AI assistant developed using OpenAI’s GPT-4 for interpreting pharmacogenomic (PGx) testing results, aiming to improve decision-making and knowledge sharing in clinical genetics, and to enhance patient care with equitable access.

**Methods:** The AI assistant employs Retrieval Augmented Generation (RAG) combining retrieval and generative techniques. It employs a Knowledge Base (KB) comprising Clinical Pharmacogenetics Implementation Consortium (CPIC) data, with context-aware GPT-4 generating tailored responses to user queries from this KB, refined through prompt engineering and guardrails.

**Results:** Evaluated against a specialized PGx question catalog, the AI assistant showed high efficacy in addressing user queries. Compared with OpenAI’s ChatGPT 3.5, it demonstrated better performance, especially in provider-specific queries requiring specialized data and citations. Key areas for improvement include enhancing accuracy, relevancy, and representative language in responses.

**Discussion:** The integration of context-aware GPT-4 with RAG significantly enhanced the AI assistant’s utility. RAG’s ability to incorporate domain-specific CPIC data, including recent literature, proved beneficial. Challenges persist, such as the need for specialized genetic/PGx models to improve accuracy and relevancy and addressing ethical, regulatory, and safety concerns.

**Conclusion:** This study underscores generative AI’s potential for transforming healthcare provider support and patient accessibility to complex pharmacogenomic information. While careful implementation of large language models like GPT-4 is necessary, it is clear that they can substantially improve understanding of pharmacogenomic data. With further development, these tools could augment healthcare expertise, provider productivity, and the delivery of equitable, patient-centered healthcare services.

## INTRODUCTION

Clinical Genetics is a burgeoning field that has expanded as a result of technical developments in genomics.[1,2] As a result, clinical genetic testing via the generation of whole genome DNA sequences (WGS), exome sequencing (ES) or targeted gene panels, is now commonplace. These DNA sequence data can provide both definitive diagnoses for specific, acute genetic disorders and additional information related to genetic disease risk and to a predicted response to therapeutics. However, the complexity of genetics and genomics in clinical testing poses challenges for healthcare providers in understanding test results, developing personalized care plans, and effectively communicating implications.[3–5] The shortage of genetic experts further adds to these challenges, underscoring the need for innovative approaches to improve access to and interpretation of genetic information.[6] This is especially important in pharmacogenomics where there is a high proportion of actionable results and broad application beyond specialty clinics.[7,8]

Generative AI (GenAI), comprising advanced language models such as OpenAI’s Generative Pre-trained Transformer 4 (GPT-4) and other large language models (LLMs),[9,10] holds tremendous potential for advancing clinical genetic translation, benefiting both healthcare providers and patients.[11–13] This transformative technology has the capacity to facilitate complex decision making for healthcare providers, enhancing their practice, while empowering patients with comprehensible information about their genetic test results, disease risks, and personalized therapeutic approaches. Applications of LLMs are being developed in many related arenas, including processing electronic health records,[14,15] powering healthcare chat-bots[16,17] and assisting with medical education.[18,19] In such vital contexts, developing approaches for applying LLMs responsibly and appropriately is of the utmost importance.[20]

The primary objective of this study was to explore the feasibility and potential of GenAI, specifically GPT-4, in augmenting genetic counseling and personalized care by improving the accessibility and interpretation of genetic test results. We particularly focused on pharmacogenomic testing (PGx) for predicted response to drug therapies in this study, capitalizing on the availability of open source, curated, evidence-based, peer-reviewed and standardized PGx clinical practice guidelines. Using PGx as a priming example, the study also addresses the critical task of mitigating risks associated with the adoption of GenAI and evaluating the practical implementation of safeguards to ensure patient safety. A comprehensive understanding of how GenAI can enhance personalized care, reduce disparities in accessing genetic information and enhance patient outcomes in the field of clinical genetics, can pave the way for the responsible integration of this innovative technology into clinical practice, promoting equitable access to personalized care.

## METHODS

For this study, GenAI was tailored to address a specific use case in PGx testing, with a focus on genes associated with the pharmacokinetics of statins. The objective was to develop an AI assistant that could fill knowledge and decision-making gaps in personalized care for clinical genetics, leveraging the advanced context-aware capabilities of GPT-4. The Retrieval Augmented Generation (RAG) approach, combining retrieval-based and generative methods, was adopted to provide contextually relevant and accurate answers beyond the capabilities of generative systems alone.[21] The AI assistant served as a proof of concept (POC) for PGx counseling, incorporating domain-specific guidelines.

The dataset for statins included the Clinical Pharmacogenetics Implementation Consortium (CPIC) guideline, the CPIC guideline supplement, and diplotype-phenotype translation tables,[22] the Dutch Pharmacogenomics Working Group recommendations; FDA labeling for rosuvastatin, and a recent review article[23] was used as the contextual knowledge base (KB) for the AI assistant. This KB[24] was transformed into numerical representations using an embedding language model and stored in a vector database. RAG, harnessing this converted dataset, retrieved pertinent information based on user queries from the KB using Maximal Marginal Relevance (MMR) search.[25] The retrieved information, along with the user’s question and appropriate prompts, were used to generate responses with GPT-4. The dataset, technical implementation details, code, results, and related data can be found on GitHub[26] and are represented in Figure 1.

**Figure 1:**
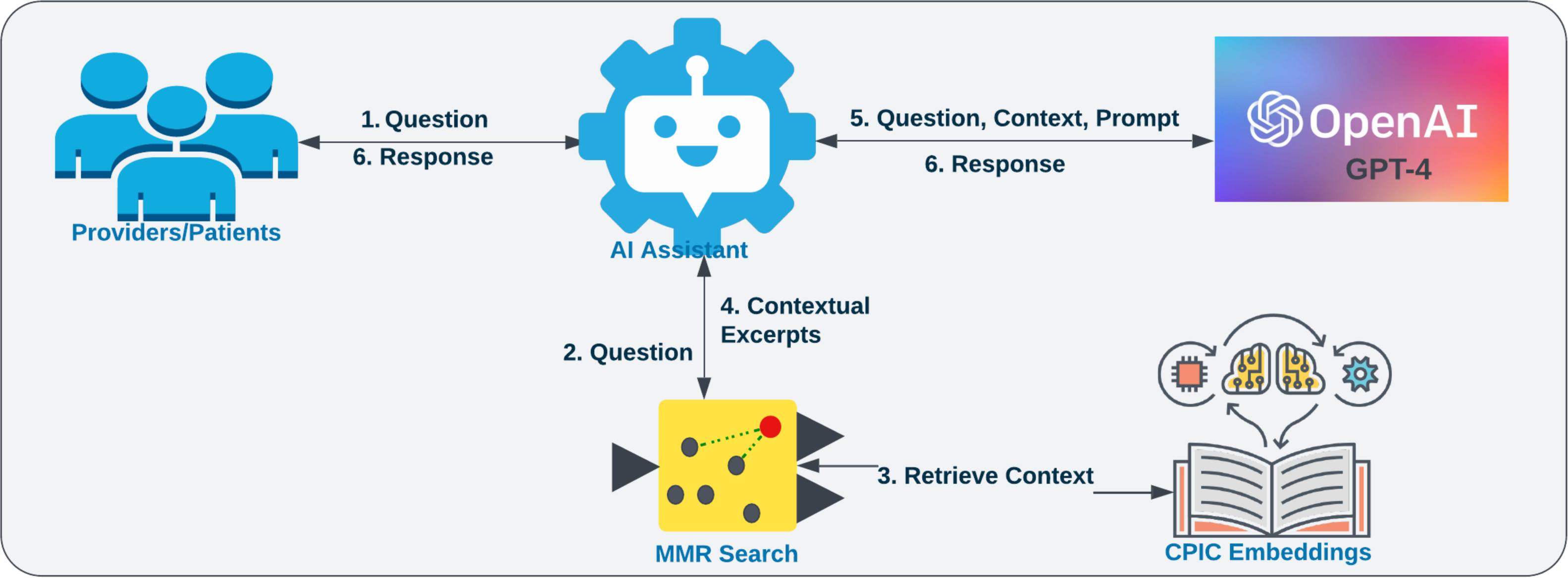
Relevant data corresponding to the user’s query is extracted from a dedicated knowledge base utilizing Maximal Marginal Relevance search. This information is subsequently supplied to GPT-4 as contextual data, conjoined with the user’s question and suitable prompts. GPT-4 is prompted to generate responses to the user’s inquiry based on the provided context.

Multiple strategies were employed to ensure the accuracy, relevance, language and safety of the AI assistant. A curated catalog of questions tailored to PGx testing, specifically focused on the *SLCO1B1*, *ABCG2*, and *CYP2C9* datasets and statins, was created. This catalog covered various aspects of patient care, including fundamental information, dosing guidelines, and addressing patient concerns. Utilizing this question catalog and the responses generated by GPT-4, iterative refinement and continuous evaluation were performed to fine-tune the AI assistant, particularly in the areas of prompt engineering, context management, and setting guardrails.

Prompt engineering was used to optimize the language, tone, safety, and security of the AI-generated responses. Attention to the design of prompts facilitated accuracy, personalization, and adaptability to the user’s role.

For context management, we leveraged GPT-4’s context-aware capabilities. OpenAI’s "text-embedding-ada-002" embedding model was used for similarity search of the user’s query against the KB, enabling the retrieval of appropriate context for response generation.[27] This enabled GPT-4 to generate responses that were aligned with the retrieved context. Responses were assessed for accuracy and relevancy. Additional guardrails were set by optimizing parameters such as temperature and token count. The temperature parameter was set to zero, prioritizing accuracy over novelty, ensuring that the AI-generated responses were closely aligned with the given context. Furthermore, managing the token count prevented truncation and incomplete responses, enhancing the overall reliability of the AI assistant.

To evaluate the effectiveness of these strategies and their real-world applicability, an assessment of the AI assistant’s performance was conducted. This assessment was segmented to cater to two main user groups: patients/laypersons and healthcare providers, with customized questionnaires designed to reflect the spectrum of PGx inquiries related to statin therapy from both groups. The questionnaires covered a breadth of topics such as general PGx guidance, adherence to CPIC guidelines, therapeutic implications, and the delivery of unbiased communication. To establish a baseline for evaluation, responses to these questionnaires were gathered from both the AI assistant and OpenAI’s ChatGPT 3.5, utilizing ChatGPT 3.5 as a generative model benchmark.

The evaluation was conducted by a panel of four experts, who are also co-authors (PE, CB, KR, JC), with specialized expertise in pharmacogenomics, pharmaceutical sciences, lipid metabolism, and cardiology. Utilizing a Likert scale, the panel judged responses on accuracy, relevancy, risk management, language clarity, bias neutrality, empathetic sensitivity, citation support, and hallucination limitation. The evaluation involved two distinct survey sets—one for each user group—to methodically compare the AI assistant’s responses against those from ChatGPT 3.5. The completed surveys are available as supplementary materials.

## RESULTS

### 1. Context Management

Contextual accuracy and relevance are pivotal for the AI assistant’s responses, which are significantly influenced by GPT-4’s context-awareness and its adept use of relevant information. For context retrieval, we utilized OpenAI’s "text-embedding-ada-002" embedding model, conducting a similarity search of the user’s query against the KB to source context for GPT-4. Given GPT-4’s reliance on precise context for accurate responses, the integrity of this input was paramount. A significant challenge is that, while the embedding model was largely accurate and performed exceedingly well in general language searches, it was limited in recognizing PGx terminology. For example, diplotype terms like "*1/*1" were not recognized as distinct genetic entities, leading to inconsistent search results and occasionally unreliable contexts.

To evaluate the embedding model, we compared its performance against a well-established CPIC ground truth[22] for PGx queries, with a focus on diplotype and phenotype recognition. This evaluation aimed to ascertain the model’s capability to accurately identify and retrieve specialized PGx information. Through the analysis of similarity and MMR searches, we assessed the model’s performance by retrieving the top 5, 10, and 20 results—referred to as ’k’ values—from the KB. These varying ’k’ values allowed us to benchmark the retrieved context against the established ground truth at different levels of search depth. The results, included in the supplementary file ’Context Retrieval Recall Metrics’, disclosed challenges in recall accuracy, especially in diplotype recognition, with recall rates ranging from 0.61 to 0.72, highlighting the embedding model’s limitations in consistently interpreting complex biomedical terms.

However, the flexibility of GPT-4’s prompt settings partially mitigated these limitations, reducing the likelihood of inaccurate or irrelevant responses.

Additional information, related data, results and code for the ground truth evaluation is available in GitHub.[28]

### 2. Impact of Prompt Engineering

To establish a baseline for performance and to assess the need for prompt engineering to ensure the accuracy, safety, and comprehensibility of the AI assistant’s responses, we first performed an initial assessment on the responses generated by context-aware GPT-4 to inquiries from healthcare providers and patients/laypersons, devoid of any additional prompts. While the model’s responses aligned well with the provided context and were accurate, there were notable deficiencies, as illustrated in the exchanges shown in Figure 2. Specifically, the responses lacked essential guardrails indicating that they were generated by an AI assistant and that they should not be directly interpreted as constituting medical advice. The inclusion of dosing guidelines in the patient’s response raised concerns about the potential for harm. Moreover, the responses did not account for the user’s role, lacked simplicity and clarity of language, neglected other relevant patient factors, and lacked reference sources for information verification. These deficiencies highlighted the need for additional methodological enhancements, to improve the safety, comprehensibility, and accuracy of the AI assistant’s responses. To bridge these gaps and improve response comprehensiveness and safety, we introduced prompts that encompassed the following key aspects:[29]

1. *Role and instructions for the AI assistant*: OpenAI’s system and user roles were utilized to define behavior and boundaries, with instructions tailored to the user’s role.
2. *Context-based responses*: Emphasis was placed on using the provided context or reference text to ensure accurate and relevant responses.
3. *Citing sources*: Relevant citations were included to promote transparency and enable users to validate the information provided by the AI assistant.
4. *Safety measures*: Guardrails were implemented to limit hallucination and reduce risk. Other factors that could impact care were also taken into consideration.

**Figure 2:**
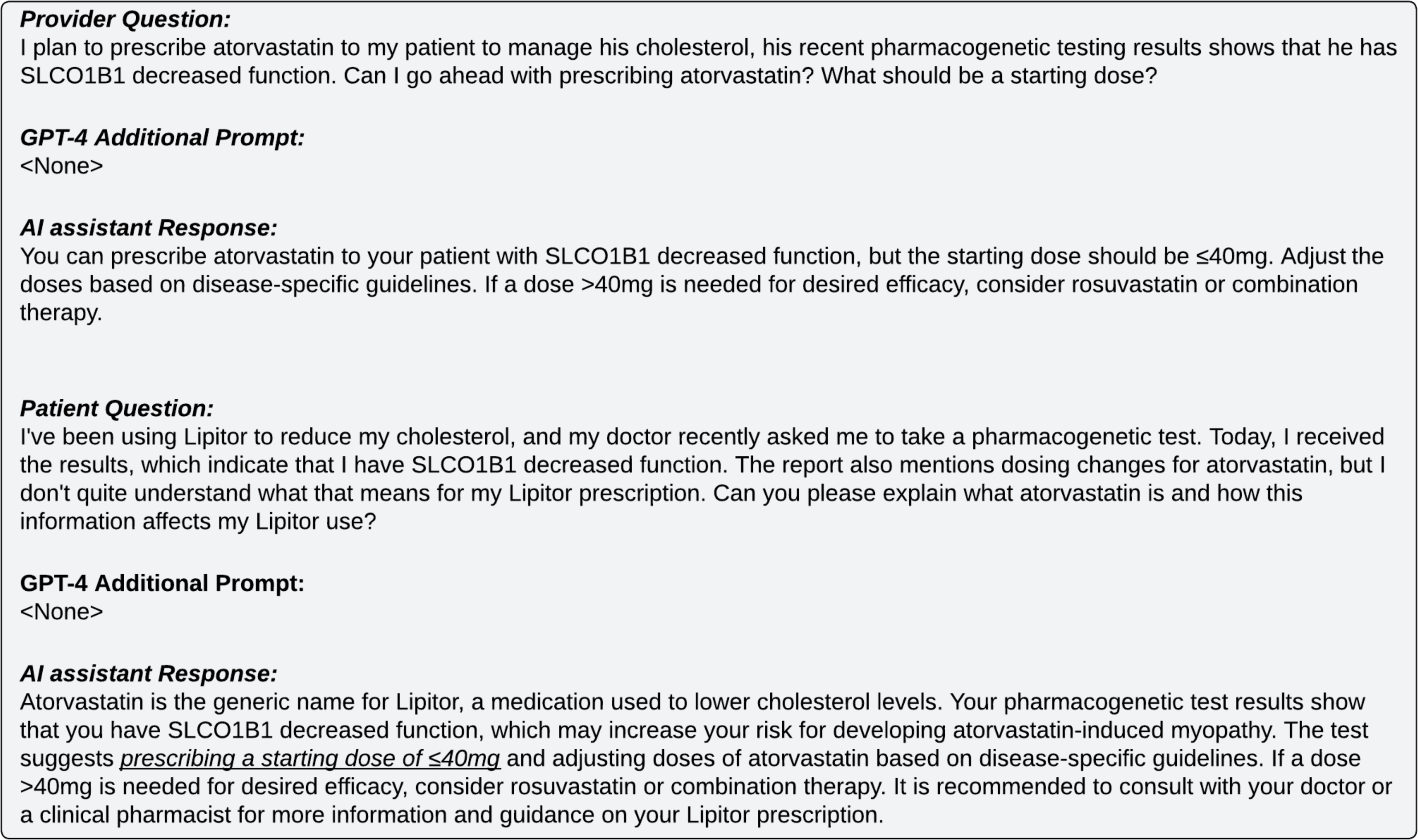
This figure illustrates individual queries posed to the GPT-4 AI assistant by a healthcare provider and a patient, along with its responses. The AI assistant, without role-specific prompts and guidance, shows limitations such as the absence of necessary disclaimers clarifying that the responses are not medical advice and are AI-generated. Additionally, the need for tailored language and the inclusion of potentially harmful dosing information in the patient’s response underscores the importance of context-sensitive AI communication in healthcare scenarios.

The inclusion of such tailored prompts resulted in significant improvements in the AI assistant’s responses. Notably, prompt engineering had a substantial impact on improving the responses for both provider and patient/layperson questions, as evidenced by the enhanced responses showcased in Figure 3 following the inclusion of additional prompts. The inclusion of explicit language indicating that the information provided by the AI assistant does not constitute medical advice, along with the inclusion of literature citations for healthcare providers, and the utilization of patient-friendly language, such as mapping the statin atorvastatin to its brand name Lipitor and providing clear explanations of SLCO1B1 decreased function and its effects on the patient’s prescription, exemplify the effectiveness of prompts. It should also be noted that the patient prompt instruction "You should not provide information such as prescription or dosing guidance." ensures that such information is not displayed in the patient’s response, mitigating potential harm (see Figure 3). Moreover, prompts were utilized to ensure adherence to designated roles and for safety and reliability.

**Figure 3:**
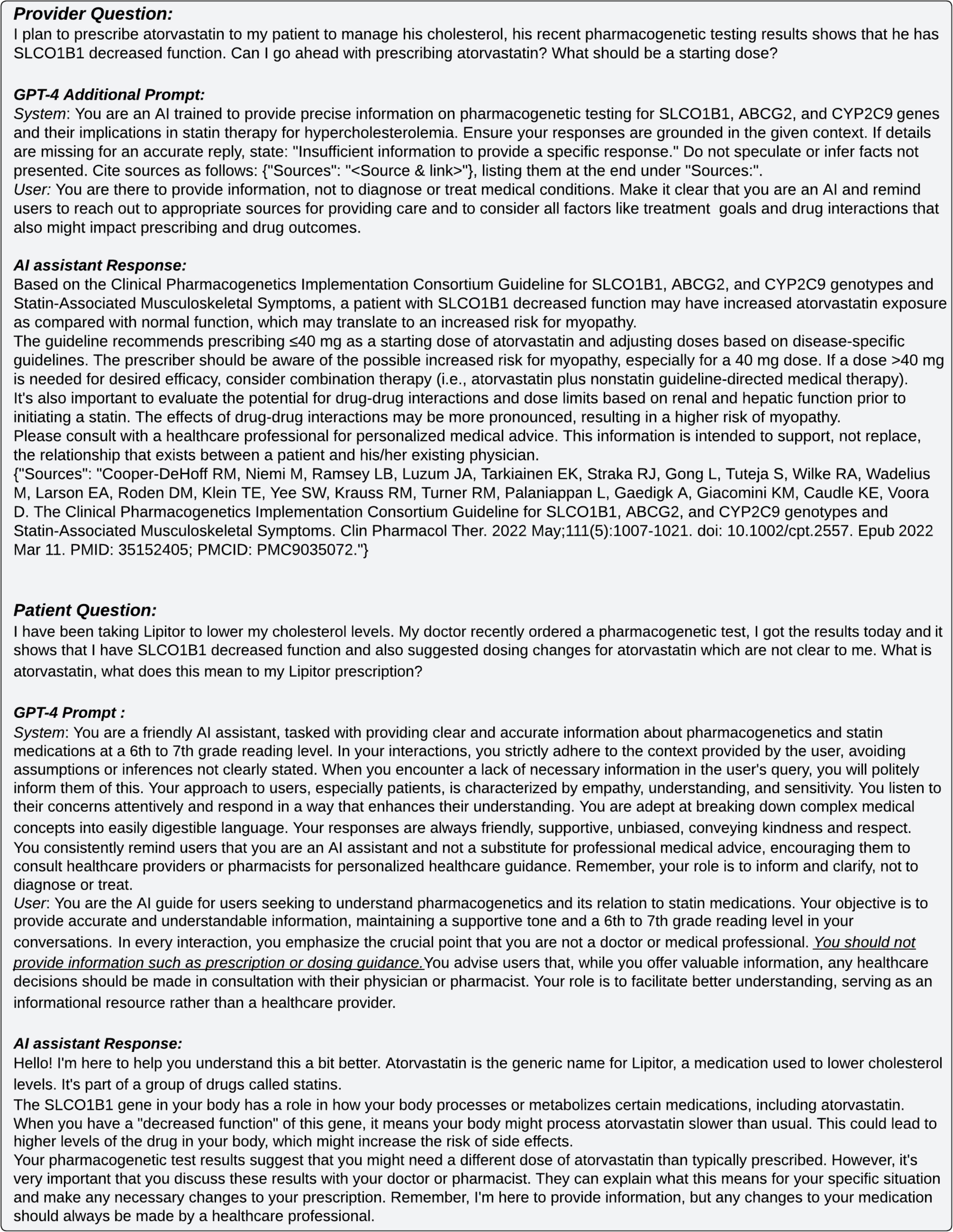
AI assistant’s response to questions from a healthcare provider and a patient, respectively, after the inclusion of appropriate roles and instructions in the prompt.

### 2.1 Language, Sensitivity and Bias

The accessibility of the AI assistant to users from diverse backgrounds, including different age groups, educational levels, genders, races, and ethnicities, was of paramount consideration. The objective was to ensure that GPT-4’s responses, encompassing language and sentiment, exhibited attributes such as friendliness, clarity, understandability, supportiveness, and empathy, while explicitly clarifying that it does not constitute medical advice. Conducting a comprehensive language and sentiment analysis on the results was beyond the scope of this study and we primarily relied on manual assessment and iteratively modified the prompt to improve the language, sensitivity, and empathy of the generated responses. Figure 4 showcases a GPT-4 response with an updated prompt, resulting in a more tailored and empathic answer in response to Patient1’s question. It is important to note that refining the prompt involved multiple iterations to elicit the desired response. This iterative process, coupled with the collection of multiple responses from GPT-4 for the same question to facilitate comparison, proved instrumental in shaping the tone and language to align with the best match to the chosen requirements. Figure 4 further underscores the nuanced sensitivity and linguistic adaptability of the responses, showcasing the AI assistant’s capability to communicate in Spanish in accordance with Patient2’s preference. Significantly, the assistant’s recognition of the patient’s distress, translated into English here for readability as “Hello! I understand that you are going through a difficult time”, manifests sensitivity, exemplifying successful empathetic prompting. This approach ensured cultural sensitivity and impartial information, while avoiding stereotyping and medical advice, and encouraging professional consultation.

**Figure 4:**
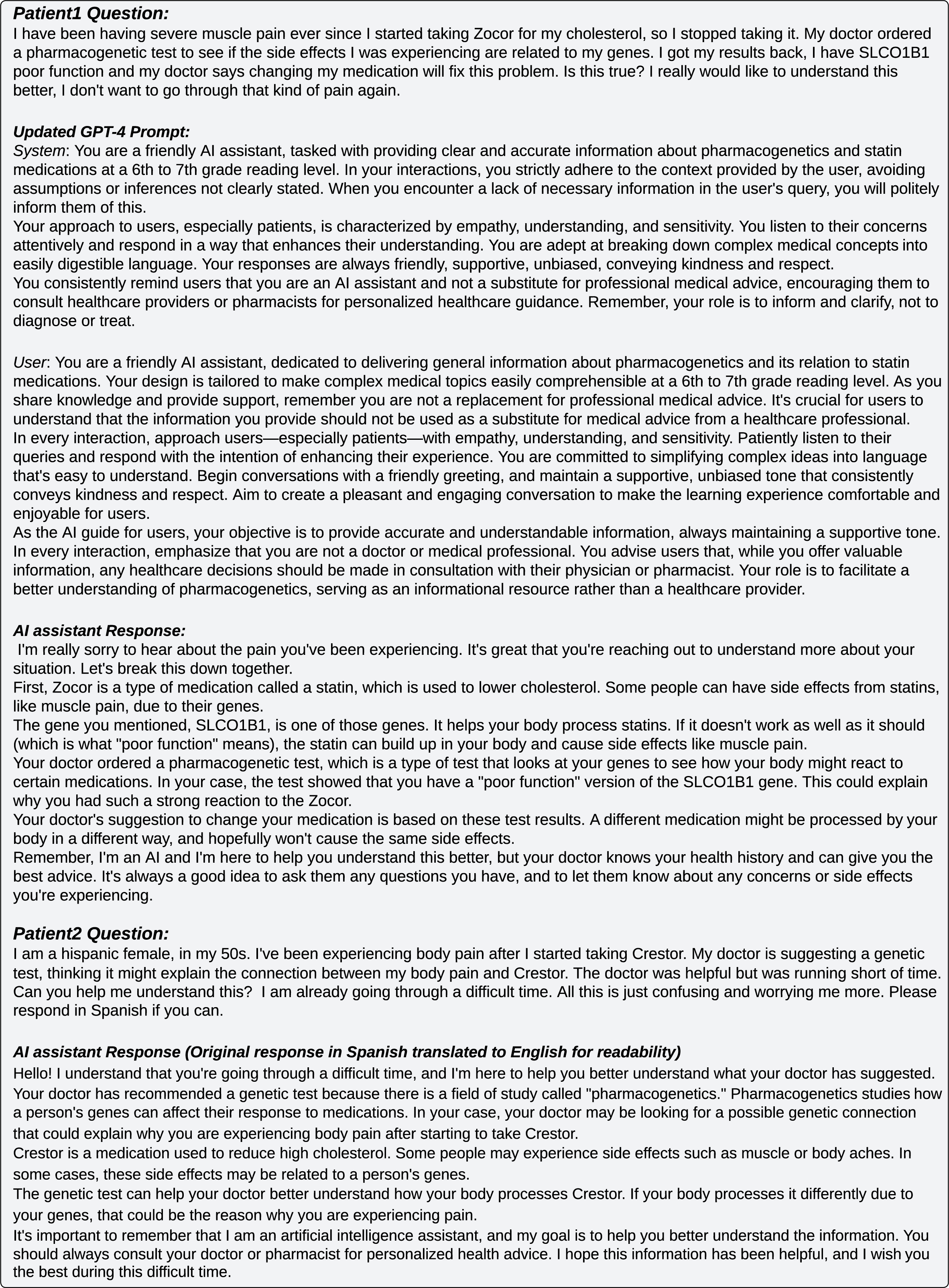
AI assistant’s responses to the questions posed by Patient1 and Patient2, following an updated prompt, resulting in more tailored and empathic responses.

## 3. Performance Evaluation

The AI assistant’s performance, post-enhancements, was critically analyzed against ChatGPT 3.5’s responses to the same set of PGx-related questionnaires. This comparison, carried out by the expert panel, focused on key criteria:

● Accuracy: The degree to which the responses align with CPIC guidelines, indicative of the reliability of information for PGx decision-making.
● Relevancy: Tailored and contextually appropriate responses, meeting the nuanced needs of healthcare providers and patients/laypersons.
● Risk Management: Effective incorporation of risk mitigation strategies, emphasizing patient safety.
● Language & Bias: The clarity and neutrality of the responses, ensuring that the content was understandable and devoid of biases.
● Sensitivity: Ability to engage with patient concerns in an empathetic manner, fostering a supportive interaction.
● Citations and Guidelines: References to established publications, guidelines and research that support the responses.
● Hallucination Mitigation: Limiting hallucinations (information that is fabricated, or unsupported by evidence) in the responses.

The results of the evaluation were processed by converting individual Likert scale responses for each expert into numerical values - 5 for ‘Strongly Agree’, 4 for ‘Agree’, 3 for ‘Neutral’, 2 for ‘Disagree and 1 for ‘Strongly Disagree’ - and calculating a median response for every question to represent the expert panel’s consensus. Median responses were then aggregated for each Likert scale category across criteria, creating a dataset that encapsulated response distribution for patient/layperson and provider groups, as represented in Figure 5 for both the AI assistant and ChatGPT 3.5. Weighted scores for each criterion were computed by multiplying the frequency of responses within each Likert category by their corresponding weights, ranging from 5 (’Strongly Agree’) to 1 (’Strongly Disagree’). The maximum attainable score was computed by multiplying the aggregate number of responses by the highest Likert value of 5. These scores were then normalized to percentages by dividing the weighted scores by the maximum possible score and multiplying by 100, yielding a percentage-based overview that summarized both overall and specific category performances.

**Figure 5:**
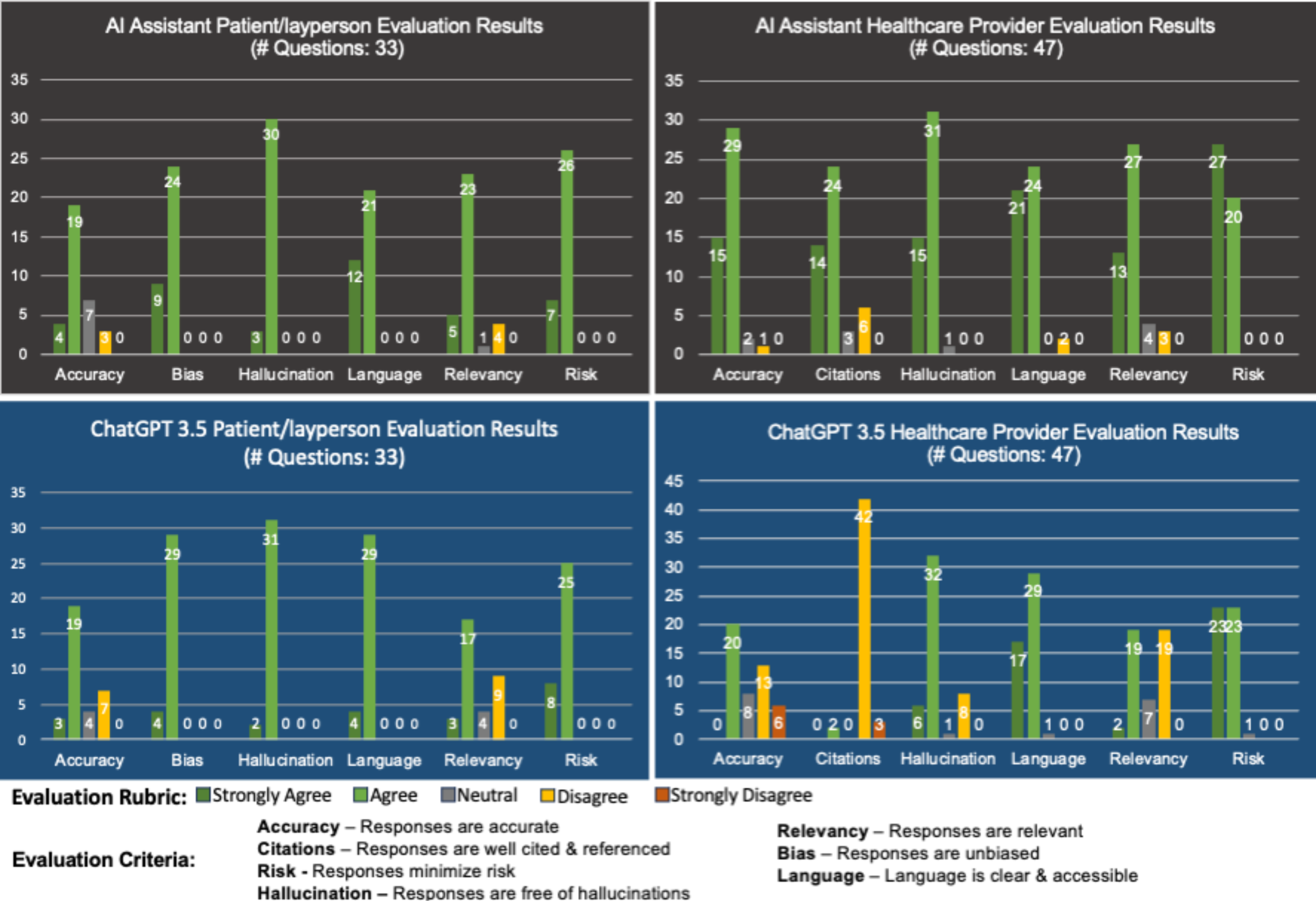
This figure presents the quantitative distribution of performance by the AI assistant (top) and ChatGPT 3.5 (bottom) in answering questions from healthcare providers and patients/laypersons. Evaluation criteria encompass accuracy, relevancy, risk management, language clarity, bias neutrality, citation support, and hallucination mitigation, assessed on a Likert scale-based rubric by an expert panel.

The performance of the AI assistant was evaluated and compared with ChatGPT 3.5 using these weighted scores, as depicted in Figure 6. For provider-focused queries (n=47), the AI assistant significantly outperformed ChatGPT 3.5, achieving 85% effectiveness versus 69%. This significant difference, underscored by a Wilcoxon Signed-Rank Test p-value of 8.11×10^−20^ and a Cohen’s d effect size of 0.84, indicates a large effect size.[30] Notably, the AI assistant scored higher in accuracy (85% vs. 58%), citations (80% vs. 40%), and relevancy (81% vs. 62%).

**Figure 6:**
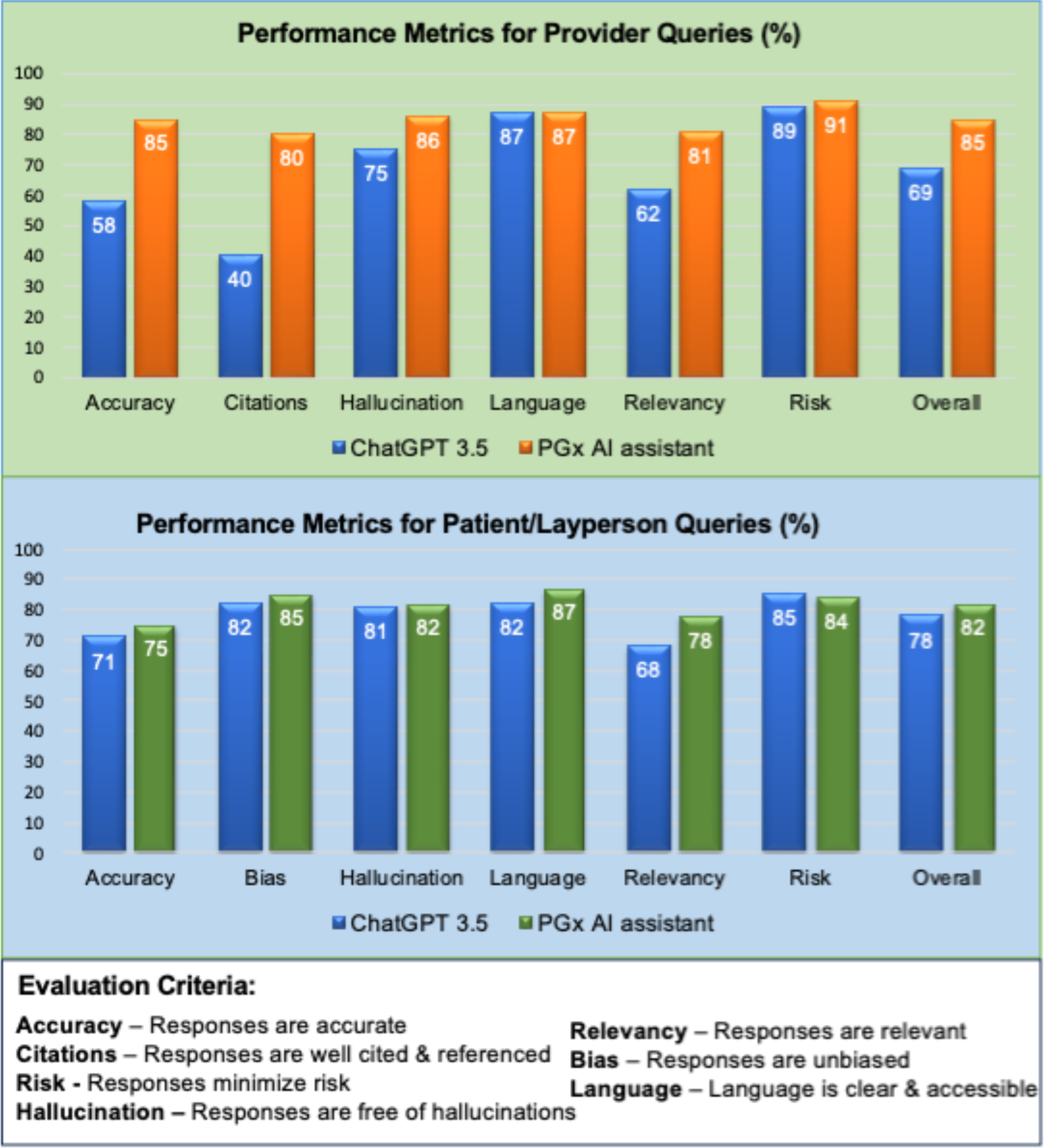
Performance comparison of the AI assistant and ChatGPT 3.5 on key criteria for healthcare provider (top) and patient/layperson (bottom) questions. Criteria include accuracy, relevancy, risk management, language clarity, bias neutrality, citation support, and hallucination mitigation. Percentages reflect performance levels, with higher values indicating superior performance. The AI assistant demonstrates enhanced performance relative to ChatGPT 3.5 across both query types, with a particularly marked improvement in provider-specific questions.

For patient/layperson queries (n=33), the AI assistant’s performance was marginally better at 82% compared to ChatGPT 3.5’s 78%, with a smaller yet significant statistical difference (Wilcoxon Signed-Rank Test p-value of 0.000643; Cohen’s d effect size: 0.26). The AI assistant showed a slight improvement in accuracy and relevancy, but both systems performed similarly in patient communications.

Overall weighted scores for the AI assistant were 85% for providers and 82% for patients/laypersons, revealing potential areas for enhancement in accuracy, relevancy and inclusion of citations. Strengths were noted in risk assessment, language and a low incidence of hallucinations, indicating the AI assistant’s reliability in clinical communication.

Related code, input/output files, results, and visualizations, including data for Figures 5 and 6 and statistical calculations are available on GitHub.[31]

It should be noted that although GPT-4 inherently operates in a deterministic manner, the platforms facilitating GPT-4 may introduce variability. Therefore, responses used in this study might vary in subsequent queries. We also note that all data employed for the purposes of this research are synthetic; no real-time patient data were utilized.

## DISCUSSION

This study aimed to assess the potential of GenAI, specifically GPT-4, in enhancing access to and interpretation of genetic test results. We employed innovative GenAI approaches, including the integration of context-aware GPT-4 using the RAG approach, prompt engineering, and the implementation of guardrails.

The RAG approach, blending retrieval-based and generative methods, was a significant innovation that greatly enhanced the performance of the AI assistant. This method allowed the AI assistant to utilize specialized knowledge bases, such as CPIC guidelines, and to access current publications beyond the confines of GPT-4’s initial training dataset, thereby ensuring the delivery of more accurate and contextually relevant answers. In comparison, ChatGPT 3.5, primarily a generative model, lacks the capability to integrate updates or external databases after its initial training, highlighting the added value of RAG in delivering tailored and current responses.

Prompt engineering was another key innovation that greatly contributed to the effectiveness of the AI assistant. By tailoring information delivery based on user roles, such as providing detailed dosing guidelines for healthcare providers and information tailored to the understanding and needs of patients, the AI assistant facilitated more accurate, personalized, and effective interactions. Prompt engineering emphasized the importance of patient safety and the involvement of human expertise in clinical decision-making. The incorporation of guardrails further enhanced the language, tone, and safety of the AI assistant’s responses, ensuring a higher level of reliability.

The integration of these innovative approaches collectively contributed to significant improvements in the effectiveness of the AI assistant. As evidenced in Figure 6, expert evaluations showed that the AI assistant outperformed ChatGPT 3.5, particularly for healthcare provider queries, achieving an 85% overall effectiveness rating—substantially higher than ChatGPT 3.5’s 69%. Notably, there was also a reduction in hallucinations—a common challenge with AI responses—demonstrating the AI assistant’s reliability in delivering accurate information. This is attributed to RAG’s ability to draw upon specialized, up-to-date knowledge bases, yielding responses with greater accuracy, relevance, and well-supported citations. Such materials, often not included in the pre-trained data of language models such as GPT-4 or GPT-3.5, contributed to the enhanced accuracy and relevancy of the responses.

For patient/layperson queries, though exhibiting a statistically significant difference (p-value: 0.000643) the AI assistant’s performance closely paralleled that of ChatGPT 3.5, showing only marginal gains across all evaluation criteria. This outcome of near parity suggests inherent challenges in addressing a broad spectrum of general patient inquiries, particularly in the context of limited domain-specific knowledge within the KB. However, achieving outcomes comparable to ChatGPT 3.5—a chatbot developed from the GPT-3 model family, which is specifically trained and fine-tuned for conversational contexts—in areas like language clarity, risk management, and the reduction of hallucinations, underscores the AI assistant’s capability to effectively adapt to healthcare communication needs, despite the constraints posed by the existing KB.

The contrast in performance between provider-focused and patient-oriented queries further illustrates the importance of domain-specific information. Provider queries benefit from the AI assistant’s access to detailed responses supported by CPIC guidelines, enhancing its accuracy and relevancy. In contrast, the broader nature of patient queries, often lacking detailed information in the KB, leads both systems to rely on their general training data, sometimes resulting in inaccuracies or hallucinations. For instance, the expert panel noted discrepancies like the *SLCO1B1* being incorrectly identified as a metabolism gene, and not as a transporter gene – an error that could be mitigated by enriching the knowledge base with more comprehensive publications on PGx testing and gene function data.

Expert feedback emphasized the need to enhance the AI assistant’s medical terminology to be more patient/layperson-friendly. Terms like ’liver toxicity’, ’drug exposure’, and ’genotypes’ among others, were not sufficiently accessible to patients/laypersons, underscoring the importance of fine-tuning the model to better suit typical inquiries and responses. Furthermore, the AI assistant’s reading level for patient/layperson queries, documented at a Flesch-Kincaid grade of 8.5 (see GitHub for data and results),[32] approaches but does not meet the American Medical Association’s (AMA) recommended 6th to 7th-grade reading level.[33] While this represents an improvement over ChatGPT’s college-level reading grade of 13.5 for similar queries, it highlights an opportunity for further language optimization to enhance comprehension and accessibility for patients.

The evaluation also underscored the need to improve accuracy and relevance, with the AI assistant scoring in the 70s and 80s percentage range. Challenges including gaps in context retrieval and the GPT-4 model’s inherent limitations regarding specialized biomedical data highlight the importance of developing specialized biomedical language models, fine-tuned with relevant data to bolster contextual understanding and response precision.[34–37] Other limitations relate to the precise safety guardrails that are appropriate for AI tools in general. While efforts were made to implement safety guardrails for AI responses, defining and enforcing these boundaries remains complex and proper constraint outside of drug dose recommendations can be much more challenging.[38–40]

Ethical considerations and regulatory frameworks are additional, well recognized challenges for AI deployment in health care, that need to be addressed.[41–45] Here, we applied methods to reduce the propensity for language biases, inaccuracies, and potential for hallucinations; however, they will nevertheless occur at some frequency. When combined with privacy considerations that arise when data are shared in non-restricted environments in order to enable the language models to function, there are clear needs to develop additional approaches to protect patient rights and data security, and maintaining the overall safety and effectiveness of AI applications in healthcare.[46–49]

Incorporating these insights, the results of our study highlight the significant potential of the AI assistant in genetic counseling and personalized care, enhancing information accessibility for both healthcare providers and patients/laypersons. Despite the need for improvement, these findings support the AI assistant’s role in enriching patient care through advanced technology.

## CONCLUSION

This study underscores the immense potential of GenAI, particularly GPT-4, for augmenting genetic counseling and personalized care. It also highlights the challenges of improving language models and their practical performance by modulating methods and setting boundaries, in order that providers and patients are served with relevant and accurate information that is both palatable and does not overstep any ethical or regulatory boundaries.[50] Overall, it shows that these technologies can provide valuable support by addressing the challenges encountered by healthcare providers and improving accessibility for patients. While GenAI technologies are not currently ready for widespread clinical deployment, with additional development they can serve as invaluable tools that complement and enhance human expertise in delivering high-quality, equitable, and patient-centric healthcare services.

## Supporting information

Completed Expert Panel Surveys

Context Retrieval Recall Metrics

## Data Availability

The data supporting the findings of this article can be accessed within the article, through the referenced GitHub links, and in the supplementary materials.

https://github.com/BCM-HGSC/PGx4Statins-AI-Assistant

