## Supplementary material for "Empowering Personalized Pharmacogenomics with Generative AI Solutions": Context Retrieval Recall Metrics

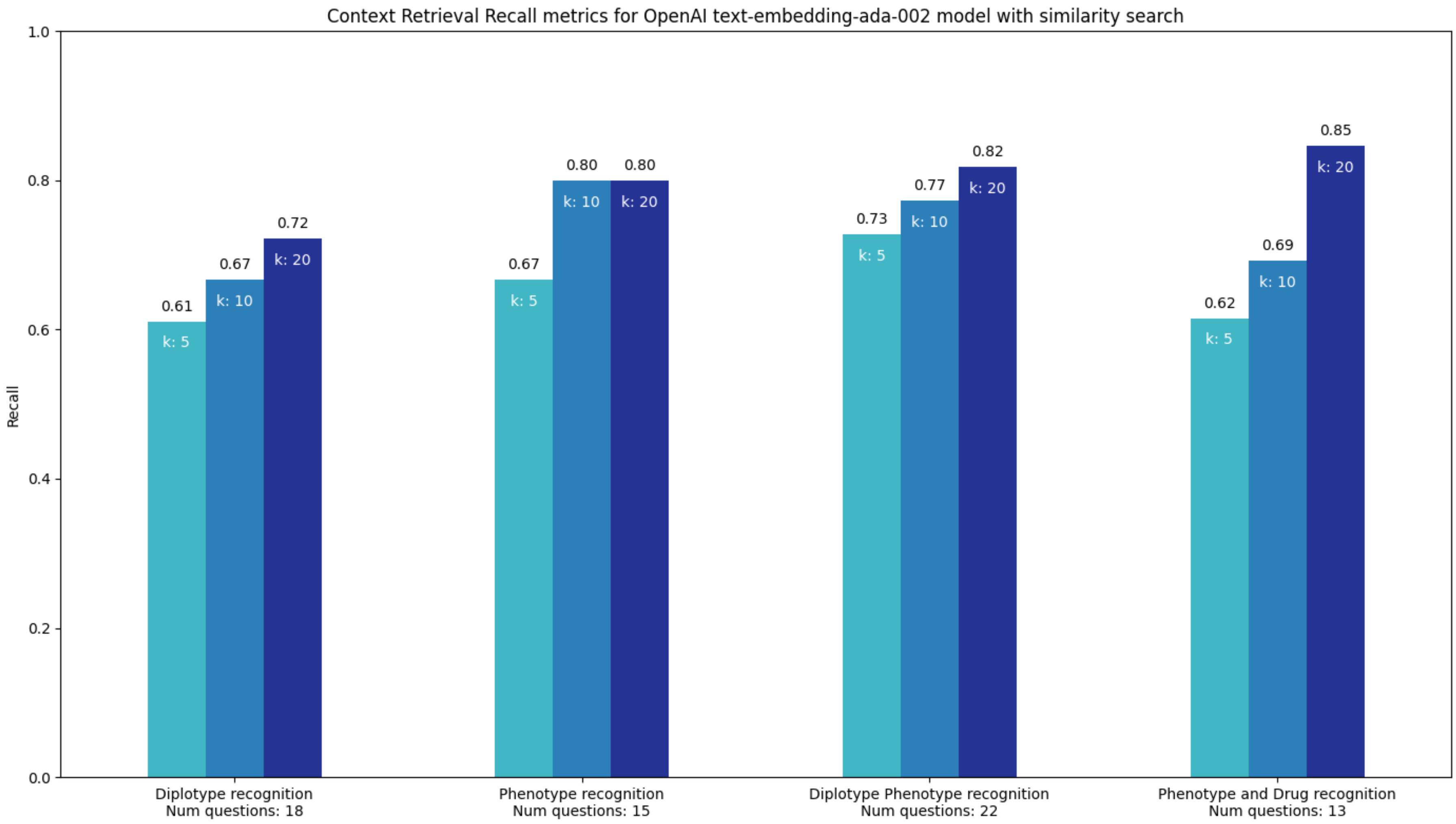

Recall Metrics for Context Retrieval. This figure illustrates the recall metrics of the OpenAI text-embedding-ada-002 model for diplotype, phenotype, and combined phenotype-drug recognition, using similarity search. It displays recall values at different retrieval depths, corresponding to the top 5, 10, and 20 results - referred to as 'k' values - from the PGx KB. The figure highlights the model's accuracy in identifying and extracting pertinent information from the PGx KB.

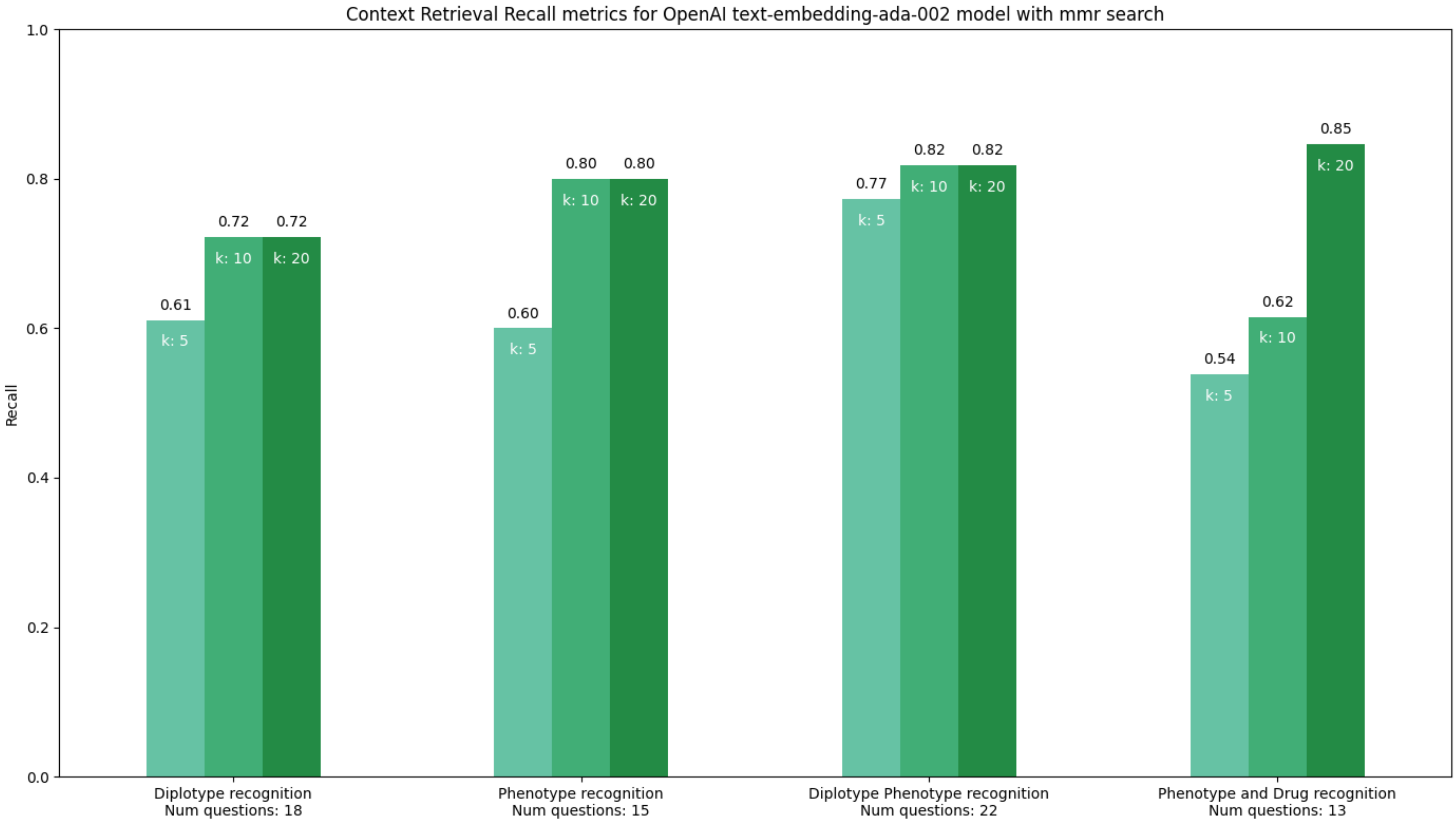

Recall Metrics for Context Retrieval. This figure illustrates the recall metrics of the OpenAI text-embedding-ada-002 model for diplotype, phenotype, and combined phenotype-drug recognition, using MMR search. It displays recall values at different retrieval depths, corresponding to the top 5, 10, and 20 results - referred to as 'k' values - from the PGx KB. The figure highlights the model's accuracy in identifying and extracting pertinent information from the PGx KB.
